# First laboratory-confirmed human case of infection with influenza A(H5N2) virus reported in Mexico

**DOI:** 10.1101/2024.08.15.24311897

**Authors:** Joel Armando Vázquez-Pérez, Claudia Wong-Arámbula, Mario Solís-Hernández, Eduardo Becerril-Vargas, Gisela Barrera-Badillo, Víctor Hugo Ahumada-Topete, Santiago Avila-Rios, Rogelio Pérez-Padilla, Fidencio Mejía-Nepomuceno, Enrique Mendoza-Ramírez, Marisol Karina Rocha-Martinez, Carlos Javier Alcazar-Ramiro, Alfredo Cruz, Joaquin Zúñiga, Karolina Bozena-Piekarska, Dayanira Sarith Arellano-Suarez, María Natividad Cruz-Ortiz, Tatiana Ernestina Núñez-García, Eréndira Molina-Gómez, Laura Adriana Flores-Cisneros, Rodrigo Aparicio-Antonio, Abril Rodríguez-Maldonado, Magaly Landa-Flores, Armando García-López, Jorge Membrillo-Hernández, Gabriel García-Rodríguez, Herlinda García-Lozano, Irma López-Martínez, Ruth P. González-Sánchez, Gustavo Reyes-Terán, Carmen M. Hernández-Cárdenas

## Abstract

In late April, 2024, we detected a non-subtypeable influenza virus in a hospitalized patient, resident of the Metropolitan area of Mexico City. Our results using 5 complete viral segments M, NS, NA, NP and HA, showed identity of 99% with avian influenza A(H5N2) from Texcoco, Mexico of 2024. This case is the first with direct evidence of human infection caused by the H5N2 influenza virus, relationship with severity remain unknow.

## Introduction

Avian influenza viruses have been implicated in several infections in humans since the 9O’s(l). The main subtypes that have been causing occasionally respiratory diseases in humans are H5N1, H7N9, H5N6 (2). Additionally, avian influenza viruses like H5N1 or H7N3 have also been implicated in human infections causing conjunctivitis but not respiratory disease (3)(4). Moreover, H5N2 has been associated mostly with migratory birds and commercial and backyard poultry infections (5).

The first avian influenza H5N2 strain isolated was the A/Chick/Penn/83 (H5N2) influenza virus was originally described infecting chickens in Pennsylvania in April 1983 and subsequently this strain became virulent in 1983 (6). To date, different viral clades of H5N2 have been described in several countries including Mexico, infecting chickens and other bird species (5).

In Mexico, government records show that low pathogenic H5N2 influenza viruses were detected in 1994 in poultry in different areas of Mexico, possibly linked to migratory bird species. At the end of 1994 highly pathogenic H5N2 strains were detected in several poultry farms causing serious economic damage amongst poultry farmers (6).

In March 2024, a high pathogenicity H5N2 outbreak was identified in a poultry farm in Michoacán state. During the same month, two low pathogenicity H5N2 outbreaks were reported in backyard poultry in the State of Mexico, in the municipalities of Texcoco and Temascalapa (7).

Some reports indicated that human cases could have occurred showing serology evidence of H5N2 infection (8)(9). However, to our knowledge no evidence of human infection caused by influenza virus H5N2 has been reported until now. Here, we described the first case of respiratory disease associated with avian influenza H5N2 in humans in Mexico.

## Methodology

Demographic data, clinical symptoms, laboratory data, and outcome-related information were obtained by electronic medical records. Clinical management was performed according to the standard of care and clinical criteria by attending physicians.

### RNA extraction and sequencing

Viral RNA was extracted from 200 μl of nasopharyngeal swabs (sample INER_INF645_24), using QIAamp Viral RNA mini kit (QIAGEN). The 8 viral genome segments were amplified simultaneously and directly from the clinical sample, using MBTuni12 and MBTuni13 primers, as described elsewhere (1-4). Libraries for the 8 viral segments were generated using the reagents of the Covid-Seq kit (Illumina, San Diego, CA, USA). Libraries were sequenced on a MiSeq sequencing platform using a 2 × 150-cycle to obtain paired-end reads (Illumina, San Diego, CA, USA). The DRAGEN COVIDSeq Targeted Microbial Pipeline on BaseSpace Sequence Hub was used for the analysis, mapping, and consensus sequence obtention. Local mapping was performed using reference sequences of H5N2 from Mexico 2019.

Sequences of five genome segments (NS, M, NA, NP and HA) of the sample INER_INF645_24 were deposited in GenBank under accession no. PP886231 - PP886235 and GISAID EPI3358335-39. Sequences of avian influenza of 2024 from Mexico were deposited in GenBank under accession no PP929863- PP929894. Blast online https://blast.ncbi.nlm.nih.gov/Blast.cgi was used to assess the identities of viral consensus sequences.

### Phylogenetic analysis

To perform phylogenetic analysis, we analyzed 222 complete genome sequences of avian influenza H5N2 available on the GenBank platform from different states in Mexico (1994-2024). Sequence alignments were created using MAFFT V7 (10) and edited with MEGA 10.0 (11). A maximum likelihood tree was constructed for the whole genome sequence using MEGA 10.0. The General Time-Reversible model was selected with five-parameter gamma-distributed rates and 1000 bootstrap replicates. Edition of the trees was made using FigTree (12).

## Results

### Clinical case

In late April, 2024, the laboratory of Molecular Biology at the National Institute for Respiratory Diseases Ismael Cosío Villegas (INER) in Mexico City reported, after confirmation by the National Institute of Diagnostics and Epidemiological Reference (InDRE) which is part of Ministry of Health of Mexico, that a nasopharyngeal sample from a man in their 60s tested positive by real-time reverse transcription-polymerase chain reaction (RT-PCR) for avian influenza A(H5N2) virus infection. INER and InDRE confirmed the H5N2 infection by RT-PCR and sequencing. Patient attended to the emergency room of the INER experiencing low back pain, diarrhea, nausea and low oxygenation rate of 80%, and was admitted for observation. He had 5 points in the Glasgow neurological scale, an arterial blood pH of 7.018, a PaCO_2_ of 43.4 mmHg, PaO_2_ of 63.5 mmHg, SaO_2_ of 77.8%, HCO3’ of 39.9 mEq/L, BE −18.2 mMol/L, lactate of 4.3 mMol/L, and a glucose of 65mg/dl. Prior informed consent, the patient was placed in invasive mechanical ventilation, and treated with IV norepinephrine, bicarbonate and volume replacement for shock and severe acidosis through a central venous catheter. Blood analysis revealed normal leucocytes, 8.41, neutrophils 8.07, lymphocytes 2, monocytes 1 (10^3^mm^3^), hemoglobin 10.5 g/dL, hematocrit 33.4, platelets 205 per mm^3^, glucose 70mg/dl, urea 379, bun 177, creatinine 15.1. calcium 6.9, magnesium 1.9, phosphorus 11, sodium 138, potasium 3.62, glycosylated hb 6.3 UNITS. D-dimer 5251 UNITS, BNP 292 UNITS, myoglobin 713 UNITS, troponin 63.8 UNITS, procalcitonin 20.85 UNITS, C reactive protein 17.4 UNITS. The patient was in peritoneal dialysis and developed peritonitis and a gram negative bacteriaemia, with a nasopharyngeal exudate positive for influenza A. A bilateral pleural effusion was observed in the chest roentgenogram. Patient did not refer traveling or exposure to poultry or domesticated animals including birds.

After 20h of evolution, the patient’s clinical status deteriorated to fatal outcome. Progressed with more ventilatory requirements, persistent hypoxemia (SaO2 49%) despite a FIO2 of 1.0, and increase of vasopressors, without reaching optimal perfusion.

### Molecular Characterization of influenza A H5N2 virus

From 160,315 total reads obtained after sequencing of sample INER_INF645_24, we obtained 11,139 (7%) unique influenza-specific reads. Using a *de novo* assembly pipeline, we obtained 5 complete segments: segment 8 (NS), 7 (M), 6 (NA), 5 (NP) and 4 (HA). The mean coverage depth was 834X, 773X, 43X, 33X and 50X respectively (GenBank accession number PP886231 - PP886235 and GISAID EPI3358335-39).

The identity of the influenza viral consensus was determined showing 97-98% with sequences of avian H5N2 from 2019 of Central Mexico (Influenza A virus (A/chicken/Queretaro/CPA-04673-l/2019(H5N2)). GenBank accession number MZ565372-MZ565375 and MZS564800 (Figure 1-3).

**Figure 1.**
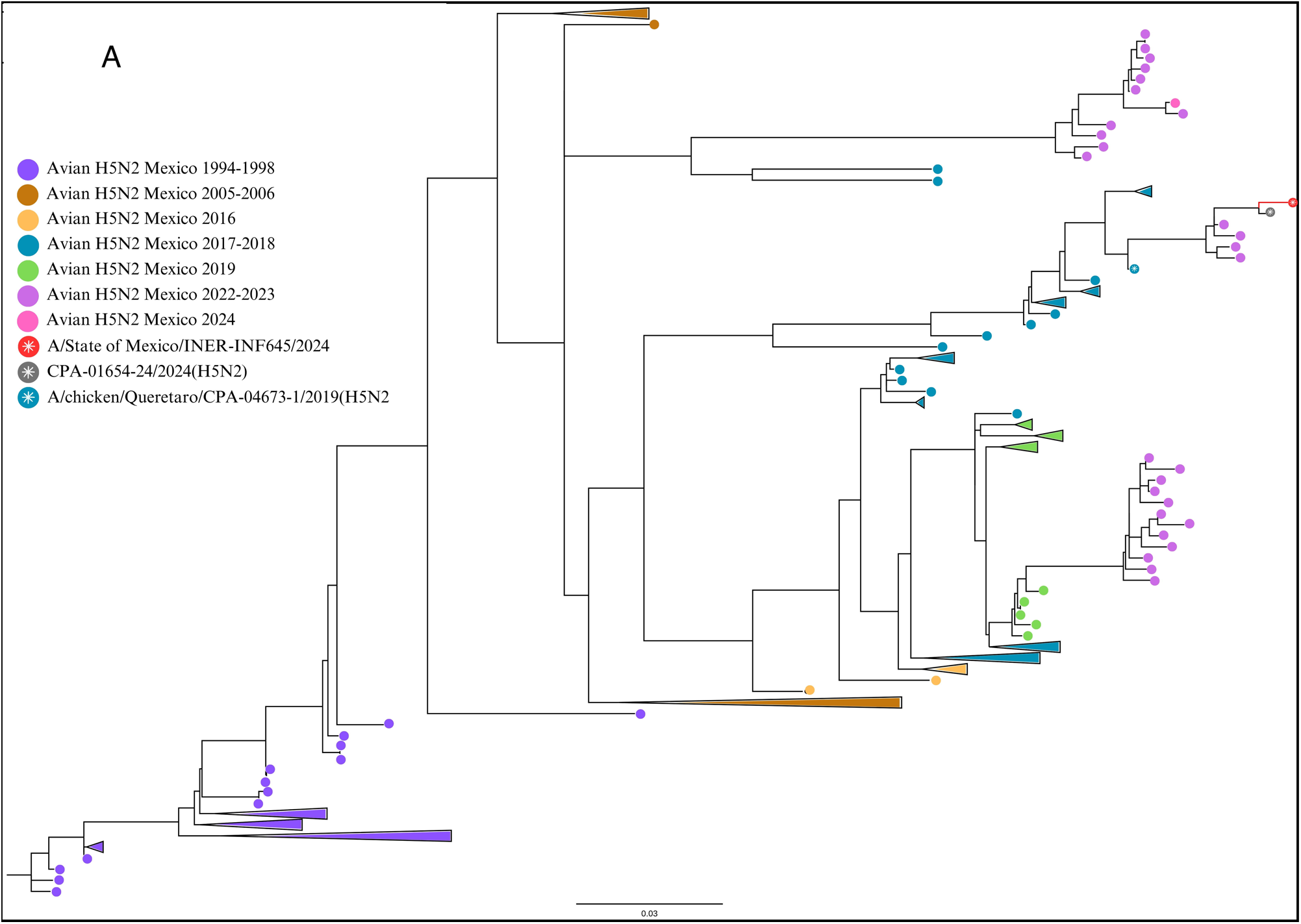
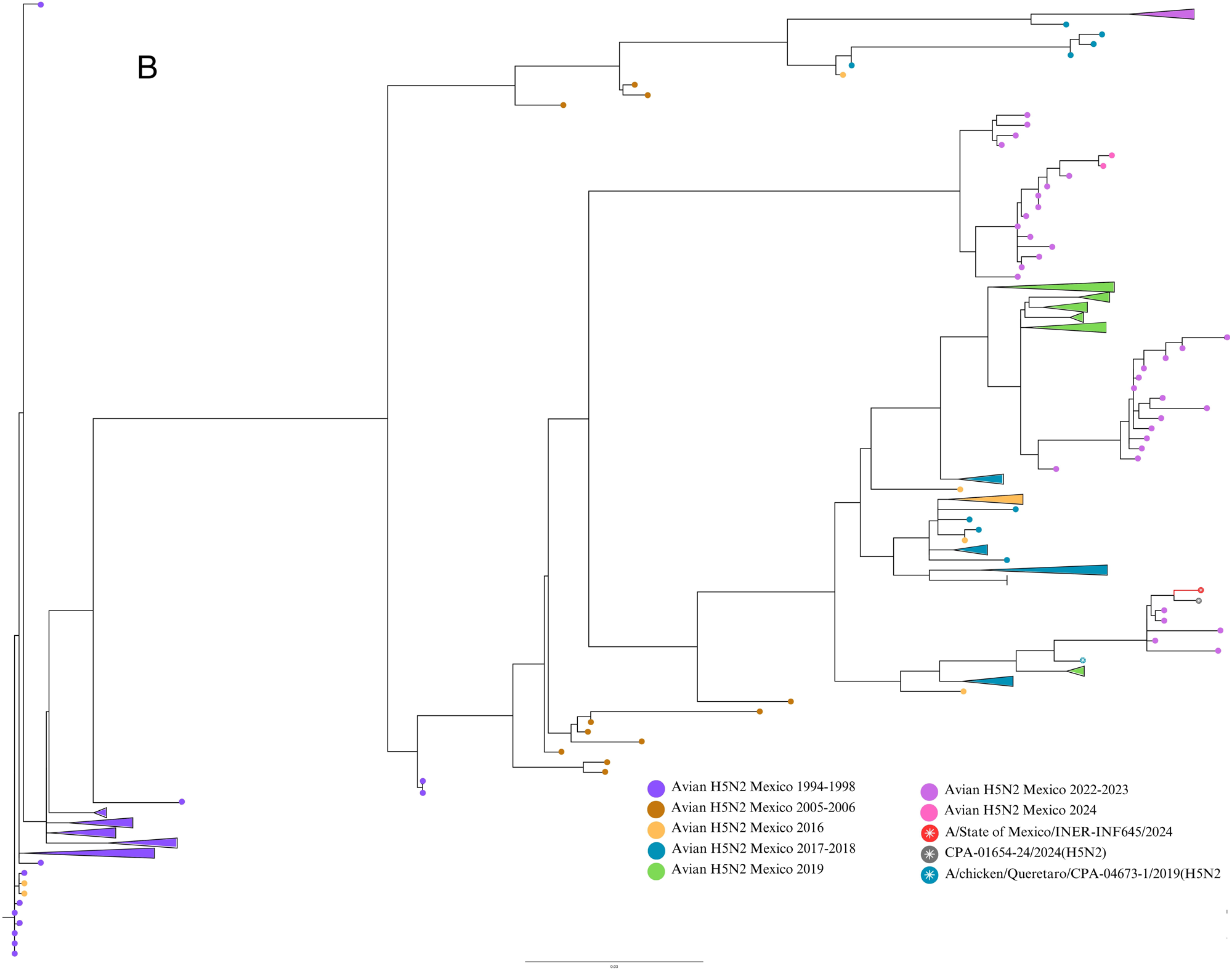
Maximum likelihood (ML) phylogenetic trees for HA (A) and NA (B) influenza H5N2 genetic segments. ML trees from 222 avian influenza H5N2 viruses registered in GenBank were produced with 1,000 bootstrap replicates. The 2024 human sequence from Mexico is included. Node with sequences from Queretaro 2019 and Texcoco 2024 are colored in red. Avian sequences from different years are indicated with brackets. Scale bar indicates nucleotide substitutions per site.

**Figure 2.**
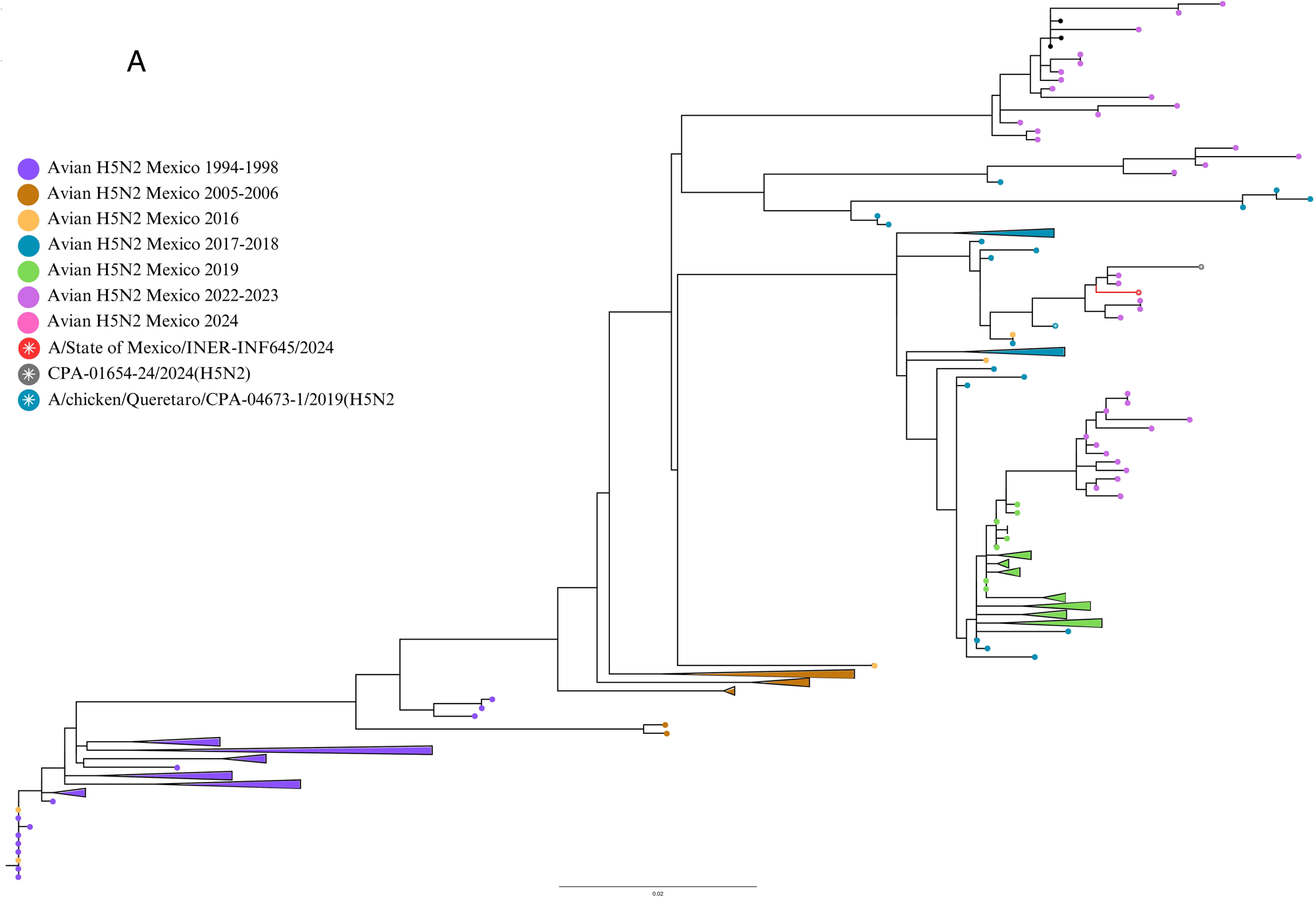
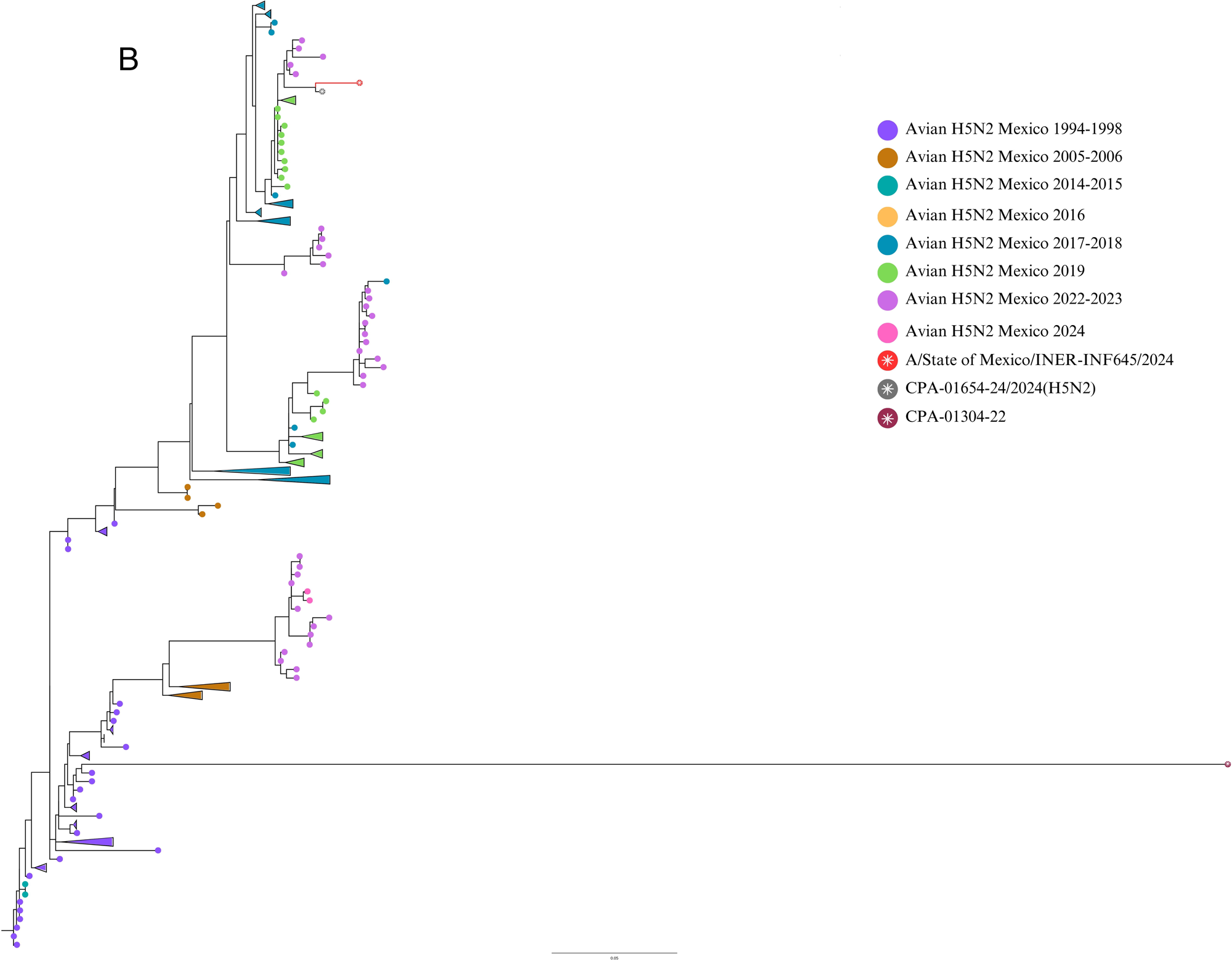
Maximum likelihood (ML) phylogenetic trees for M (A), NS (B) influenza H5N2 genetic segments. ML trees from 222 avian influenza H5N2 viruses registered in GenBank were produced with 1,000 bootstrap replicates. The 2024 human sequence from Mexico is included. Node with sequences from Queretaro 2019 and Texcoco 2024 are colored in red. Avian sequences from different years are indicated with brackets. Scale bar indicates nucleotide substitutions per site.

**Figure 3.**
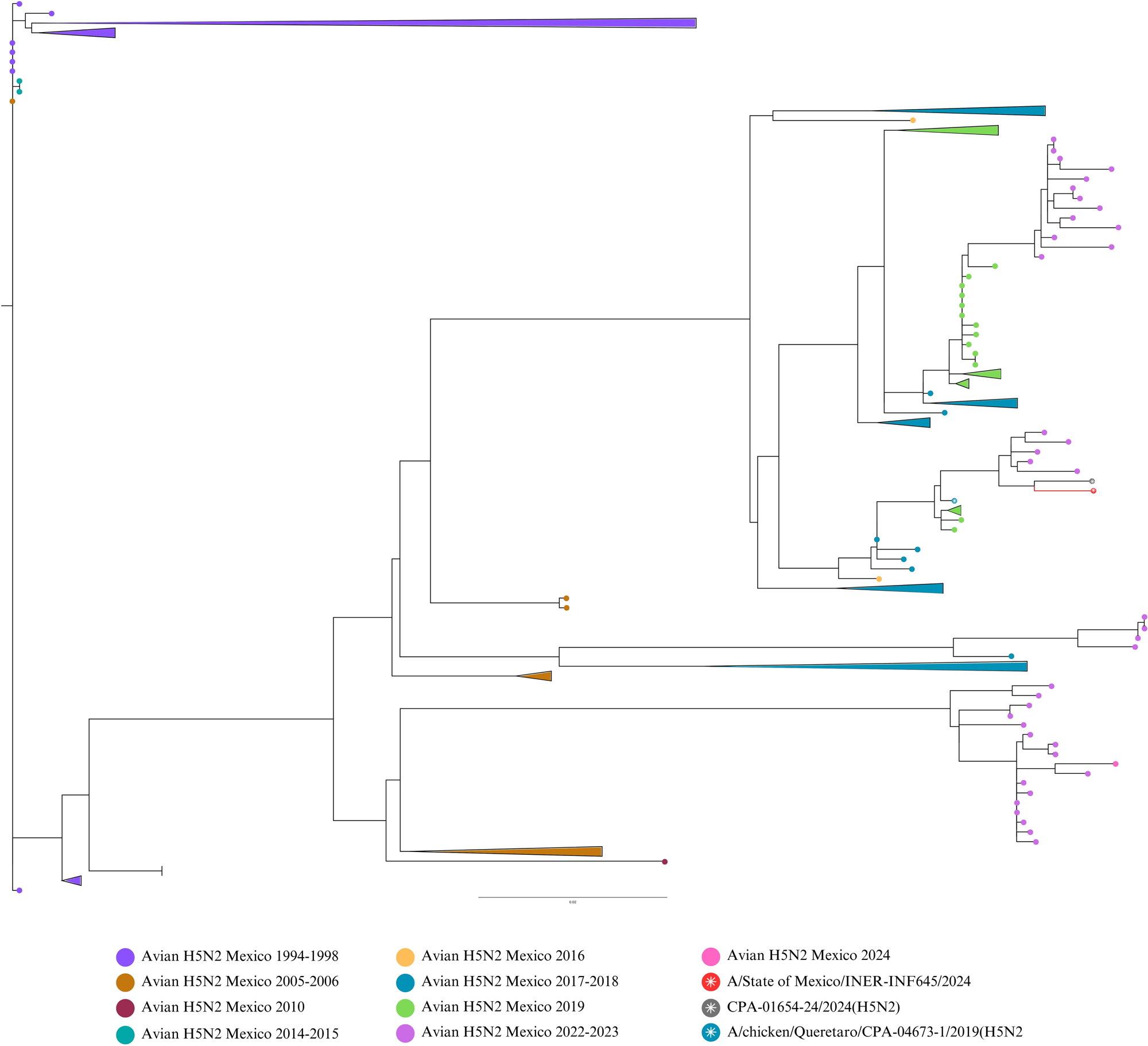
Maximum likelihood (ML) phylogenetic trees for NP influenza H5N2 genetic segment. ML trees from 222 avian influenza H5N2 viruses registered in GenBank were produced with 1,000 bootstrap replicates. The 2024 human sequence from Mexico is included. Node with sequences from Queretaro 2019 and Texcoco 2024 are colored in red. Avian sequences from different years are indicated with brackets. Scale bar indicates nucleotide substitutions per site.

Through mapping with reference (H5N2_Mex_2019, MZ565371), we obtained a partial sequence of segment 3 (PA), with 80% coverage.

Using more recent H5N2 sequences from Mexico (2022, 2023 and 2024), we performed a complete phylogenetic analysis. The sequences of segments M, NA, NP and HA clustered with one sequence of 2019 (A/chicken/Queretaro/CPA-04673-1/2019(H5N2) and with sequences of 2022, 2023 and 2024. Likely, the isolate of 2019 was the predecessor of the isolates clustered together with the human case of the present study. The closest identity (99%) was for a 2024 avian sequence from State of Mexico (GenBank accession number PP929863-PP929894). This sequence is related with high disease burden and mortality in backyard farms in Texcoco, a locality in the State of Mexico near to the residence of the patient of the present study. Moreover, there were only 3 non-synonymous mutation differences between the two isolates, human and avian from Texcoco 2024. E489A in HA, G114R in NS1 and T433P in NP. The NS segment showed a different pattern, clustering with a separate group of samples from 2022-2023 and showed lower identity with the 2019 isolate.

The multi-basic amino acids at the cleavage site in HA, showed three basic amino acids: PQKRKR/G. According to previous studies (5), the observed cleavage site sequence would classify the studied virus as a low pathogenicity isolate.

We were not able to detect characteristic mutations of highly pathogenic influenza viruses, including E627K in PB2 of H5N1 and H7N7 viruses (13), because of the low yield of amplification and sequencing of polymerases genes. H255Y in NA, that confers resistance to oseltamivir in H5N1 viruses, was not present in this isolate.

## Discussion

Detection and molecular characterization of influenza virus H5N2 in a respiratory sample, confirmed the first report of human infection due of this subtype in Mexico.

Molecular evidence suggests that the human isolate of this study (INER_INF645_24) and the avian isolates from 2022, 2023 y 2024, possibly derive from a common avian H5N2 ancestor from 2019 of Central Mexico (Influenza A virus (A/chicken/Queretaro/CPA-04673-l/2019(H5N2)). The observation of the highest homology (99%) of the study virus and an avian H5N2 isolate from Texcoco, State of Mexico (2024), suggests a direct relationship between these isolates. Although a direct contact of the patient in this study with poultry or other domestic animals could not be confirmed, it is plausible that this avian virus, causing high disease burden in chickens in this geographical area in 2024 could be the source of the human case described here, as human to human transmission seems unplausible.

This is the first report of a human case of influenza H5N2 infection in Mexico. Further studies are required to determine the predicted pathogenicity of the virus and to predict its capability for human-to-human transmission and potential threat to human health. Unfortunately, several comorbidities in the case described here led to a fatal outcome, but the pathogenicity of the isolate needs to be further studied.

Since no cases of H5N2 influenza in humans have been reported so far, we are unaware of the clinical outcomes that this influenza virus subtype may have in humans. Referring to the clinical manifestations associated with other currently circulating avian influenza viruses, such as H5N1, which has documented transmission to humans causing acute respiratory infections as well as mild symptoms where conjunctivitis has notably been the main manifestation in documented cases. At admission the patient was severely ill, with uremia, renal failure and important metabolic acidosis and shock state, that deteriorated progressively, and a bacteremic infection originating likely from peritonitis. It is uncertain what was the contribution of the influenza virus H5N2 to the final clinical status of the patient and also unknown how the patient acquired the influenza virus very similar to bird viruses identified in the valley of Mexico in 2022.

Dr. Vazquez-Perez is a head of Molecular Biology and Emerging Diseases Laboratory at Instituto Nacional de Enfermedades Respiratorias (INER). His research interests are the molecular epidemiology and pathogenesis of respiratory viruses, and Dr. Hernandez-Cardenas is a General Director of Instituto Nacional de Enfermedades Respiratorias (INER). Her research interests are pulmonary chronic and acute diseases. Is a qualified critical care physician, a registered internist and anesthesiologist.

## Data Availability

All data produced in the present study are available upon reasonable request to the authors

https://gisaid.org/

## Acknowledgments

We thank Eduardo Márquez García from the Unidad de Biología Molecular, INER for technical assistance in Illumina sequencing. We also thank all physicians in the ICU for assistance with patient management.

This work was financially supported by Direccion General de Politicas de Investigacion en Salud (DGPIS), Grant “FPIS2024-INER-4886” to J.A.V.-P.

## Informed Consent Statement

This study was reviewed and approved by the Science, Biosecurity and Bioethics Committee of the Instituto Nacional de Enfermedades Respiratorias (protocol number B22-23). Informed consent was provided according to the Declaration of Helsinki. Written informed consent was obtained from the patients and/or from their relatives or authorized legal guardians prior to the publication of this paper.

## References

1. Beigel JH, Farrar J, Han AM, Hayden FG, Hyer R, de Jong MD, et al. Avian influenza A (H5N1) infection in humans. N Engl J Med. 29 de septiembre de 2005;353(13):1374–85.

2. Philippon DAM, Wu P, Cowling BJ, Lau EHY. Avian Influenza Human Infections at the Human-Animal Interface. J Infect Dis. 23 de julio de 2020;222(4):528–37.

3. CDC [Internet]. 2016 [citado 24 de junio de 2024]. CDC Newsroom. Disponible en: https://www.cdc.gov/media/releases/2024/p0401-avian-flu.html

4. Lopez-Martinez I, Balish A, Barrera-Badillo G, Jones J, Nunez-Garcia TE, Jang Y, et al. Highly pathogenic avian influenza A(H7N3) virus in poultry workers, Mexico, 2012. Emerg Infect Dis. 2013;19(9):1531–4.

5. Xu W, Navarro-Lopez R, Solis-Hernandez M, Liljehult-Fuentes F, Molina-Montiel M, Lagunas-Ayala M, et al. Evolutionary Dynamics of Mexican Lineage H5N2 Avian Influenza Viruses. Viruses. 3 de mayo de 2022;14(5):958.

6. Horimoto T, Rivera E, Pearson J, Senne D, Krauss S, Kawaoka Y, et al. Origin and molecular changes associated with emergence of a highly pathogenic H5N2 influenza virus in Mexico. Virology. 20 de octubre de 1995;213(l):223–30.

7. Rural S de A y D. gob.mx. [citado 24 de junio de 2024]. Registra influenza aviar AH5N2 de alta patogenicidad una unidad de traspatio de Michoacan. Disponible en: http://www.gob.mx/agricultura/prensa/registra-influenza-aviar-ah5n2-de-alta-patogenicidad-una-unidad-de-traspatio-de-michoacan?idiom=es

8. Ogata T, Yamazaki Y, Okabe N, Nakamura Y, Tashiro M, Nagata N, et al. Human H5N2 avian influenza infection in Japan and the factors associated with high H5N2-neutralizing antibody titer. J Epidemiol. 2008;18(4):160–6.

9. Yamazaki Y, Doy M, Okabe N, Yasui Y, Nakashima K, Fujieda T, et al. Serological survey of avian H5N2-subtype influenza virus infections in human populations. Arch Virol. 2009;154(3):421–7.

10. Katoh K, Misawa K, Kuma K ichi, Miyata T. MAFFT: a novel method for rapid multiple sequence alignment based on fast Fourier transform. Nucleic Acids Res. 15 de julio de 2002;30(14):3059–66.

11. Kumar S, Stecher G, Li M, Knyaz C, Tamura K. MEGA X: Molecular Evolutionary Genetics Analysis across Computing Platforms. Mol Biol Evol. 1 de junio de 2018;35(6):1547–9.

12. Rambaut A. FigTree [Internet], [citado 13 de marzo de 2024]. Disponible en: http://tree.bio.ed.ac.uk/software/figtree/

13. Liu WJ, Li J, Zou R, Pan J, Jin T, Li L, et al. Dynamic PB2-E627K substitution of influenza H7N9 virus indicates the in vivo genetic tuning and rapid host adaptation. Proc Natl Acad Sci U S A. 22 de septiembre de 2020;117(38):23807–14.

